# Analytical benchmarking of extraction-free lysis devices for molecular detection of tuberculosis

**DOI:** 10.64898/2026.07.24.26358841

**Authors:** Sonal Jain, Alexey Ball, Caitlin Anderson, Adithya Cattamanchi, Claudia M. Denkinger, Amy Steadman, Seda Yerlikaya

**Affiliations:** Department of Infectious Diseases and Tropical Medicine, Heidelberg University Hospital and Faculty of Medicine, Heidelberg University, Heidelberg 69120, Germany; Global Health Labs, Inc., Bellevue, WA 98007, USA; Center for Tuberculosis, Institute for Global Health Sciences, University of California San Francisco, San Francisco, CA 94143, USA; Division of Pulmonary Diseases and Critical Care Medicine, University of California Irvine, Irvine, CA 92697, USA; German Center for Infection Research, Partner Site Heidelberg University Hospital, Heidelberg 69120, Germany

## Abstract

Sample preparation remains a barrier for decentralized, swab-based molecular testing of tuberculosis (TB). Extraction-free workflows offer a simpler alternative, but systematic benchmarking against standard methods is lacking. We evaluated five novel lysis devices, BLINK Shaker Prototype, nPOC-BB, SPS-1, Truelyse, and Thermolyse, against a heat- and bead-beating reference method using contrived *M. tuberculosis*-spiked tongue and sputum swabs. The primary outcome was lysis efficiency, measured as the relative DNA recovery compared with the reference workflow. Secondary outcomes included nuclease inactivation, biosafety, and usability. In the reference buffer, lysis efficiencies ranged from 51–63% to 95– 154% on tongue swabs and 12–54% to 280–644% on sputum swabs across the five devices. In proprietary buffers, performance varied more widely, with lysis efficiencies of 2–4% to 64– 80% on tongue swabs and 1% to 94–398% on sputum swabs. Complete biosafety inactivation was achieved by three devices; two showed residual growth (<0.02%). Lysis efficiency of several devices met or exceeded the reference, supporting the feasibility of extraction-free workflows for TB diagnosis, with further optimization of buffer compatibility and biosafety profiles expected to enhance performance.

## Introduction

Tuberculosis (TB) remains the world’s deadliest respiratory infection, in large part because an estimated 25% of patients go undiagnosed each year,^1^ driven primarily by limited access to diagnostics. Despite World Health Organization (WHO)’s recommendations for rapid molecular testing, only 54% of individuals with TB received such a test as their initial workup in 2024,^1^ highlighting the urgent need for simpler, decentralized diagnostic approaches - a direction reinforced by the recent WHO recommendation of the first near-point-of-care TB test.^2^

Sample processing remains a principal driver of diagnostic complexity. Sputum is the gold standard sample type for TB diagnosis but is intrinsically difficult to process.^3,4^ Conventional molecular workflows require multiple steps for liquefaction, inactivation, lysis, and DNA extraction prior to detection. The lipid-rich, mycolic acid-dense cell wall of *Mycobacterium tuberculosis* (MTB) further renders it particularly resistant to heat or chemical lysis alone, historically necessitating mechanical agitation and specialized equipment that is difficult to deploy outside laboratory settings.^5–8^

Extraction-free sample preparation offers a path to simpler, more affordable testing. Swab-based sampling reduces matrix complexity and enables crude lysis without dedicated equipment.^9–11^ Tongue swabs are now WHO-recommended for adults unable to produce sputum and have proven sensitive, patient-friendly, and cost-effective.^2^ Steadman, Andama *et al*. previously demonstrated efficient MTB detection from tongue swabs using heat and bead-beating paired with manual quantitative polymerase chain reaction (qPCR), establishing a robust extraction-free reference workflow.^12^ Crucially, this swab-based, extraction-free approach has since been extended to sputum swabs, broadening its applicability. Building on this, several manufacturers have developed portable lysis devices employing diverse mechanical lysis mechanisms, some combined with proprietary buffers, that can be paired with affordable isothermal or PCR-based assays without requiring integrated extraction. Detection workflows incorporating some of these devices (Truelyse; Molbio Diagnostics, India and Thermolyse; Guangzhou Pluslife Biotech, China) have already received WHO recommendation.^2^ However, independent analytical benchmarking of lysis efficiency and biosafety across devices, against a gold-standard reference, remains limited, leaving a critical evidence gap for informing device selection and future development.

In this study, we systematically evaluated the lysis efficiency of five novel lysis devices; secondary objectives included evaluating the nuclease inactivation efficiency, biosafety, and operational usability.

## Methods

### Study design

We conducted a systematic analytical benchmarking study comparing five extraction-free lysis devices against a heat and bead-beating reference workflow. Experiments were performed under BSL-2 conditions at Global Health Labs (Seattle, USA) in accordance with institutional biosafety protocols. Tongue swab samples were collected from healthy volunteers who provided written informed consent.

### Sample collection

Tongue swab collection followed an established consensus protocol.^13^ Briefly, samples were self-collected using sterile flocked swabs (Copan FLOQSWAB 520CS01), with participants swabbing the visible anterior three-quarters of the tongue, focusing on the posterior dorsum. Participants provided up to six swabs per collection, with a maximum of two collection time points per day. Normal human pooled sputum was purchased from healthy donors (Seracare 0375-0002). Multiple stocks were pooled and subsequently aliquoted into 50 mL conical tubes to minimise sample heterogeneity. The swab head was swirled 2-3 times inside the conical tube (Figure 1).

**Figure 1:**
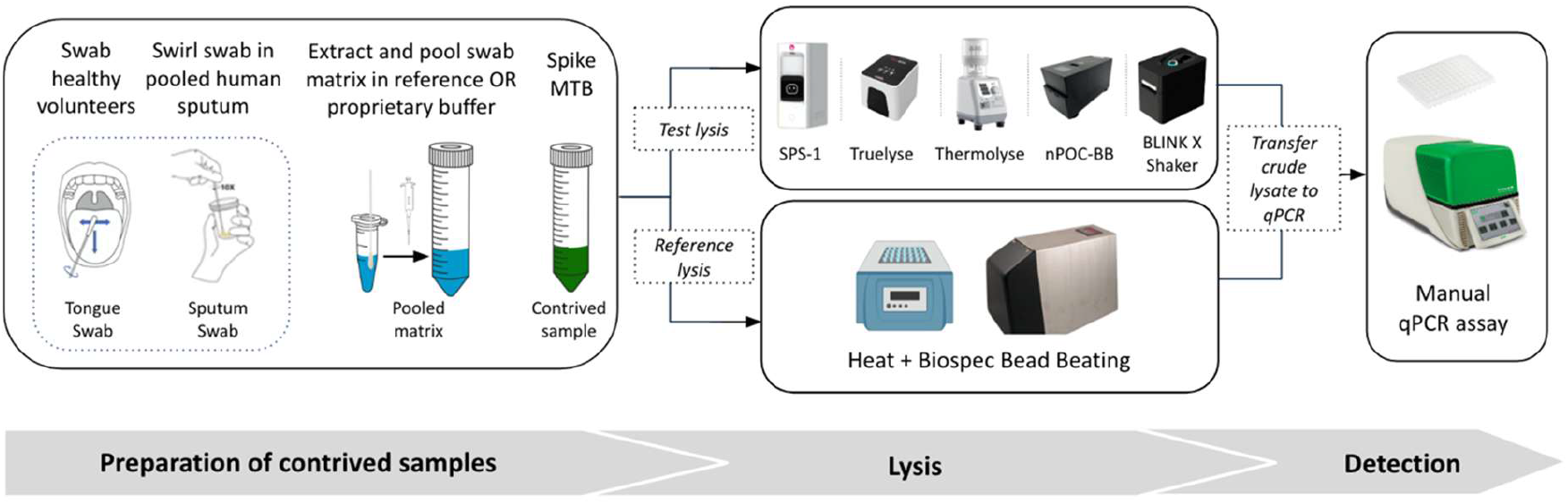
Overview of the experimental workflow for evaluating the lysis efficiency of extraction-free lysis devices against a reference workflow of heat and bead-beating in contrived tongue swabs and sputum swabs for molecular detection of MTB. Abbreviations: MTB, Mycobacterium tuberculosis; qPCR, quantitative polymerase chain reaction.

### Contrived sample preparation

Immediately after collection, swab heads from both tongue and sputum swabs were transferred into 2 mL tubes and broken at the 30 mm breakpoint. 500 µL Tris-EDTA (TE) pH 8.0, used as the reference buffer, was added to each tube. For evaluation of lysis devices with proprietary buffers, swabs from each donor were alternated between TE and the device-specific proprietary lysis buffer. Tubes containing swab heads were vortexed at maximum speed for 15 seconds to ensure proper mixing. Maximum volume of eluates (380-400 µL) from individual tubes were transferred using a micropipette into a 50 mL conical tube to generate pooled tongue swab matrix (TSM) and pooled sputum swab matrix (SSM) (Figure 1).

### MTB cell culture

0.5 mL of thawed MTB H37Ra (ATCC 25177) glycerol stock was added to a 50 mL conical tube containing 9.5 mL growth media (Middlebrook 7H9 with ADC, 0.1% Tween 80, 0.2% glycerol) pre-warmed to room temperature. Cells were expanded at 37°C on a horizontal tube roller for 5-7 days until they reached an optical density (OD) of 1-1.2, following previously described procedures.^12^

Prior to experimentation, 1-2 mL of MTB H37Ra culture was removed from 4°C storage and vortexed vigorously on high for 60 seconds to ensure thorough resuspension. A single wash step was performed by centrifuging the cells at 3000 *x g* for 2 minutes, removing the supernatant, and resuspending them in 1 mL fresh media. Cell concentrations were adjusted to an OD of 1, corresponding to ∼1-2 × 10^6^ CFU/µL. An initial dilution was prepared by transferring the cells into media containing Tween-80 to maintain monodispersity. Cells were then further diluted down to the desired concentrations by adding them directly into the pooled matrices.

### Lysis of contrived samples

#### Reference method

A previously described heat and bead-beating (BB) protocol was used as the reference method.^12^ Briefly, contrived samples were heated at 95°C for 10 minutes followed by bead-beating using the Biospec Mini-Beadbeater-16 (BioSpec 607EUR) for three minutes. Heat-only and no-treatment controls were included in each experimental run to monitor MTB H37Ra culture integrity. Heat-only controls were incubated at 95°C for 10 minutes, while the no-treatment controls underwent no lysis procedure. Based on prior optimization experiments, lysis efficiencies of <10% for the heat-only control and <2% for the no-treatment control, relative to the reference method, were considered acceptable. Biological replicates were generated by aliquoting spiked TSM or SSM into independent tubes for separate processing.

#### Index methods

A summary of the technical specifications of the index lysis devices is presented in Table 1 and serial numbers are listed in Supplementary 1, Table S1.

**Table 1:** Technical specifications and operational characteristics of the novel lysis devices. (Abbreviations: n/a, not applicable)

| Product Name | BLINK Shaker Prototype | nPOC-BB | SPS-1 | Truelyse | Thermolyse |
| --- | --- | --- | --- | --- | --- |
| Developer Name | BLINK Dx | Global Health Labs | Coyote Bioscience | Molbio Diagnostics | Guangzhou Pluslife Biotech |
| Developer Country | Germany | USA | China | India | China |
| Development stage | Prototype | Prototype | Design-locked | Design-locked | Design-locked |
| Mechanism | Bead-beating | Bead-beating | Sonication | Sonication | Vortexing |
| Shaking motion | Vertical | Horizontal | n/a | n/a | Orbital |
| Heater | n/a | 90°C | n/a | n/a | 75°C |
| Lysis vials | 2 mL screwcap cryovial | 3 mL rapid extraction tubes | Custom | Custom | 3 mL rapid extraction tubes |
| Lysis beads | 0.1mm ceramic, 600 mg | 0.1 mm glass, 150mg | n/a | Undisclosed | Custom |
| Dimensions (wxdxh in cm) | 16.2 x 10.5 x 15 | 9 x 15 x 11.4 | 22 x 12.5 x 29.8 | 12.4 x 11.5 x 10.7 | 10.7 x 14.6 x 25.1 |
| **Weight (kg)** | 3.7 | 1.7 | 5 | 1.2 | 3 |
| **Battery Powered** | No | Yes | No | Yes | No |
| **Lysis duration (minutes)** | 6 | 6 | 4 | 2 | 5 |
| **Sample capacity** | 3 | 1 | 1 | 1 | 3 |
| **Sample input volume (μL)** | 1500 | 500 | 800 | Undisclosed | 2000 |

- <u>BLINK Shaker Prototype (BLINK Dx, Germany</u>: 500 µL of the contrived sample was added to 2 mL manufacturer-supplied screw-cap tubes containing 600 mg of 0.1 mm ceramic beads. Samples were heated at 95°C for 10 minutes and then processed on the BLINK Shaker Prototype for a total of 6 minutes (three 1-minute cycles with 1-minute rests between cycles, plus a 1-minute rest at the end) at 3000 RPM.
- <u>nPOC-BB (Global Health Labs, USA)</u>: 500 µL of contrived sample was added to 2 mL tubes containing 150 mg of 0.1 mm glass beads. Tubes were processed in the nPOC-BB for a total of 6 minutes (3 minutes heating at 95°C followed by 3 minutes of shaking).
- <u>SPS-1 (Coyote Bioscience, China):</u> 800 µL of contrived sample was loaded into SPS-1 cartridges and subjected to sonication inside the device for a total of 90 seconds (two 30-second cycles with a 30-second rest between cycles).
- <u>Truelyse (Molbio Diagnostics, India)</u>: The contrived sample was added to Truelyse tubes and processed according to the device’s preprogrammed protocol.
- <u>Thermolyse (Guangzhou Pluslife Biotech, China):</u> 2 mL of contrived sample was added to Pluslife lysis tubes and processed on Thermolyse for 5 minutes (simultaneous heating and shaking at 75 °C, 3000 RPM).

### Evaluation of lysis efficiency

Lysis efficiency was evaluated across three input concentrations representing high (10^6^ colony-forming units [CFU]/mL), medium (10^4^ CFU/mL), and low (10^2^ CFU/mL) MTB levels. These concentrations were selected based on previously reported bacterial loads in clinical tongue swab samples.^12^ Following sample processing, all lysates were maintained on ice prior to qPCR.

### Quantitative PCR (qPCR) analysis

qPCR was performed using previously described primers and probes targeting the IS6110 insertion sequence and human RNaseP as internal control.^12^ 10 µL crude lysate was added to a 96-well plate containing 10 µL master mix. Samples were processed on a Bio-Rad CFX real-time PCR instrument in technical triplicates with cycling parameters described previously.^12^ IS6110 copy numbers were calculated from MTB H37Rv genomic DNA (gDNA) (ATCC 25618DQ) standard curves.

### Assessment of nuclease inactivation

To evaluate the potential for target DNA degradation, we assessed the nuclease inactivation efficiency of each lysis device in tongue swab matrices at medium cell input (10^4^ CFU/mL). Nuclease activity was evaluated post-lysis to determine the stability of the samples prior to amplification. Following the lysis step, half the samples were incubated at 37°C for 45 minutes to promote residual nuclease activity and compared to samples kept on ice, which served as a baseline. The percentage of nuclease inactivation was calculated by dividing the DNA recovery of samples incubated at 37 °C by that of samples kept on ice.

### Data analysis

Results were exported from the Bio-Rad CFX96 software to Microsoft Excel using a fixed threshold of 100,000 RFU for downstream analysis. Quantification cycle (Cq) values were averaged across three technical replicates for each experimental condition. For experiments performed with the same device, standard curves were averaged across plates and used to calculate IS6110 copy numbers. Lysis efficiency was calculated as the ratio of the measured genomes/µL for each test device to that of the reference lysis method in TE buffer, multiplied by 100.

### Biosafety verification

MTB cells were spiked into TE buffer or proprietary buffer at a concentration of 5 x 10^8^ CFU/mL and subjected to lysis using the index lysis device. Reference lysis, heat-only, and no lysis controls were also included. 200 µL lysate were plated onto Middlebrook 7H11 agar plates (Hardy Diagnostics W35), corresponding to a final concentration of ∼1 × 10^8^ CFU/plate. All conditions were tested in replicates of five, except for Thermolyse, which was evaluated in three replicates due to sample volume limitations. A no-MTB negative process control was included for each condition. Plates were incubated at 37°C and monitored for bacterial growth for up to eight weeks, with imaging performed at four weeks and visual confirmation at eight weeks. Bacterial viability was estimated based on colony counts from one quarter of the plate and normalized to the input bacterial load.

### Operational feasibility and user experience

User experience was assessed through a survey conducted at the end of the study. Each device was evaluated independently by the same three laboratory staff members who had performed the benchmarking experiments, following the manufacturer’s instructions for use (IFU), with each member blinded to others’ scores. After completing each procedure, they rated each usability parameter on a Likert scale of 1-5 (1 = Strongly Disagree to 5 = Strongly Agree) and recorded any deviations, difficulties, or notable observations.

## Results

### Heat and bead-beating in TE buffer established as the reference lysis workflow

To establish a baseline for maximum lysis efficiency, a combination of heat and bead-beating was used as the reference lysis method. In all experimental runs, lysis efficiency for heat-only controls remained below 10%, while no-treatment controls yielded less than 2% DNA recovery relative to the reference. These results established a rigorous benchmark against which all subsequent device performance data were normalised.

### Lysis efficiency is influenced by sample matrix and buffer composition

Proprietary buffers were confirmed to be compatible with downstream qPCR amplification (Supplementary 1, Figure S1) prior to testing lysis efficiency. Table 2 summarises the lysis efficiency results for all devices across MTB inputs, sample types, and buffer conditions, presented as percent recovery normalised against the reference lysis method. Key performance trends are described below for each device.

**Table 2:** Lysis efficiency (measured as percent DNA recovery) at high, medium, and low MTB input levels for each lysis device. Values are reported as mean ± standard deviation from 10 biological replicates per condition, except for Thermolyse which was tested in 5 replicates. Abbreviations: MTB, Mycobacterium tuberculosis; CFU, Colony Forming Units; TE, Tris-EDTA.

| Test device: BLINK Shaker Prototype |  |  |  |  |
| --- | --- | --- | --- | --- |
| MTB Input<br>(CFU/mL) | TE Buffer |  |  |  |
|  | Tongue Swab |  | Sputum Swab |  |
| 10<sup>6</sup> | 105% ± 6.4% |  | 106% ± 13.9% |  |
| 10<sup>4</sup> | 92% ± 3.6% |  | 84% ± 6.0% |  |
| 10<sup>2</sup> | 67.6% ± 51.6% |  | 40.6% ± 38.4% |  |
| Test device: nPOC-BB |  |  |  |  |
| MTB Input<br>(CFU/mL) | TE Buffer |  |  |  |
|  | Tongue Swab |  | Sputum Swab |  |
| 10<sup>6</sup> | 108.6% ± 8.8% |  | 136.5% ± 20.5% |  |
| 10<sup>4</sup> | 111% ± 12.9% |  | 97.9% ± 25.1% |  |
| 10<sup>2</sup> | 79% ± 78.6% |  | 72.2% ± 89.8% |  |
| Test device: SPS-1 |  |  |  |  |
| MTB input<br>concentration<br>(CFU/mL) | TE buffer |  | Proprietary Buffer |  |
|  | Tongue Swab | Sputum Swab | Tongue Swab | Sputum Swab |
| 10<sup>6</sup> | 78.6% ± 13.0% | 175.9% ± 21.9% | 28.7% ± 8.7% | 315.8% ± 40.4% |
| 10<sup>4</sup> | 92% ± 38.7% | 130.1% ± 43.8% | 31.5% ± 10.3% | 199.8% ± 62.3% |
| 10<sup>2</sup> | 153.8% ± 221.1% | 87.9% ± 77.5% | 20.8% ± 35.3% | 126.2% ± 95.6% |
| Test device: Truelyse |  |  |  |  |
| MTB Input<br>(CFU/mL) | TE buffer |  | Proprietary Buffer |  |
|  | Tongue Swab | Sputum Swab | Tongue Swab | Sputum Swab |
| 10<sup>6</sup> | 100.5% ± 5.8% | 544% ± 67.3% | 79.7% ± 9.0% | 397.9% ± 35% |
| 10<sup>4</sup> | 95% ± 13.9% | 365% ± 67.3% | 64% ± 7.3% | 93.9% ± 12.5% |
| 10<sup>2</sup> | 154.1% ± 85.3% | 280.4% ± 148% | 64.3% ± 31.8% | 131.8% ± 99.7% |

*Table 2: Lysis efficiency (measured as percent DNA recovery) at high, medium, and low MTB input levels for each lysis device.
| Test device: Thermolyse |  |  |  |  |
| --- | --- | --- | --- | --- |
| MTB Input<br>(CFU/mL) | TE buffer |  | Proprietary Buffer |  |
|  | Tongue Swab | Sputum Swab | Tongue Swab | Sputum Swab |
| 10<sup>6</sup> | 61.9% ± 8.9% | 11.6% ± 3.2% | 3.2% ± 0.3% | 1% ± 0.1% |
| 10<sup>4</sup> | 51.2% ± 3.9% | 31.9% ± 14.7% | 2% ± 0.2% | 0.6% ± 0.1% |
| 10<sup>2</sup> | 62.8% ± 8.4% | 53.9% ± 16% | 4.2% ± 3.2% | 1.5% ± 1.6% |

#### BLINK Shaker Prototype (heat + bead-beating)

The BLINK Shaker Prototype demonstrated lysis efficiency comparable to the reference method for both tongue and sputum swabs in TE buffer. For tongue swabs, DNA recovery ranged from 67.6-105%, while sputum swabs ranged from 40.6-106%, with greater variability at the lowest input.

#### nPOC-BB (heat + bead-beating)

The nPOC-BB platform yielded lysis performance comparable to the reference method across both sample matrices in TE buffer. Tongue swab recoveries ranged from 79-111%, while sputum swabs ranged from 72-137% across MTB inputs. Performance variability increased at the lowest input concentration.

#### SPS-1 (sonication)

SPS-1 lysis efficiency was equivalent to the reference method in TE buffer for both sample types, but was reduced in the proprietary buffer for tongue swabs. Lysis efficiency in TE buffer ranged from 79-154% for tongue swabs; however, use of the proprietary buffer reduced recovery to 21-32% across concentrations. For sputum swabs, DNA recovery ranged from 88-176% in TE buffer and 126-316% in the proprietary buffer.

#### Truelyse (sonication with beads)

Truelyse demonstrated matrix and buffer dependent performance. For tongue swabs, recovery was comparable to the reference ranging between 95-154% in TE buffer and 64-80% in the proprietary buffer. For sputum swabs, DNA recovery reached or exceeded the reference in TE buffer (280-544%) and proprietary buffer (94-398%).

#### Thermolyse (heat + vortex with beads)

Thermolyse performance was found to be strongly buffer-dependent. For tongue swabs, lysis efficiency was moderate relative to the reference ranging between 51-63% in TE buffer and reduced to 2-4% in the proprietary buffer. Similarly, lysis efficiency in sputum swab was 12-54% in TE buffer and reduced to 1% in the proprietary buffer relative to the reference.

### Nuclease inactivation and biosafety profiles varied across lysis devices

The bead-beating systems, BLINK Shaker Prototype and nPOC-BB, achieved 86.5% and 70.6% nuclease inactivation, respectively. Among the sonication-based systems, Truelyse and SPS-1 demonstrated 85.8% and 100% inactivation, respectively. Thermolyse, which combines simultaneous heat and vortexing, achieved complete (100%) nuclease inactivation (Supplementary 1, Figure S2).

Processing with the BLINK Shaker Prototype, nPOC-BB, and Thermolyse resulted in a complete loss of viability, with no growth observed over an eight-week incubation period (Supplementary 1, Figures S3-S9). Minimal bacterial growth was detected in samples processed using Truelyse and SPS-1 (Supplementary 1, Figures S10-S13), with <0.02% viable cells. Growth observed in control plates (heat + bead-beating and heat-only) was attributable to non-MTB contamination, as suggested by colony morphology and growth characteristics, and did not confound interpretation of MTB viability outcomes.

### Overall usability was high, with device-specific limitations identified

Usability results are summarised in Figure 2 and full parameters and scores are listed in Supplementary 2. Overall, all devices received high scores for setup and preparation. In contrast, lower scores were observed for specific sample loading and handling steps, including tube insertion in the BLINK Shaker Prototype and risk of incorrect loading in Thermolyse. Procedures and accompanying instructions were rated as easy to follow and sufficient. User interface and controls, particularly on-device display of key parameters such as target temperature, timer, and speed, were rated highly.

**Figure 2:**
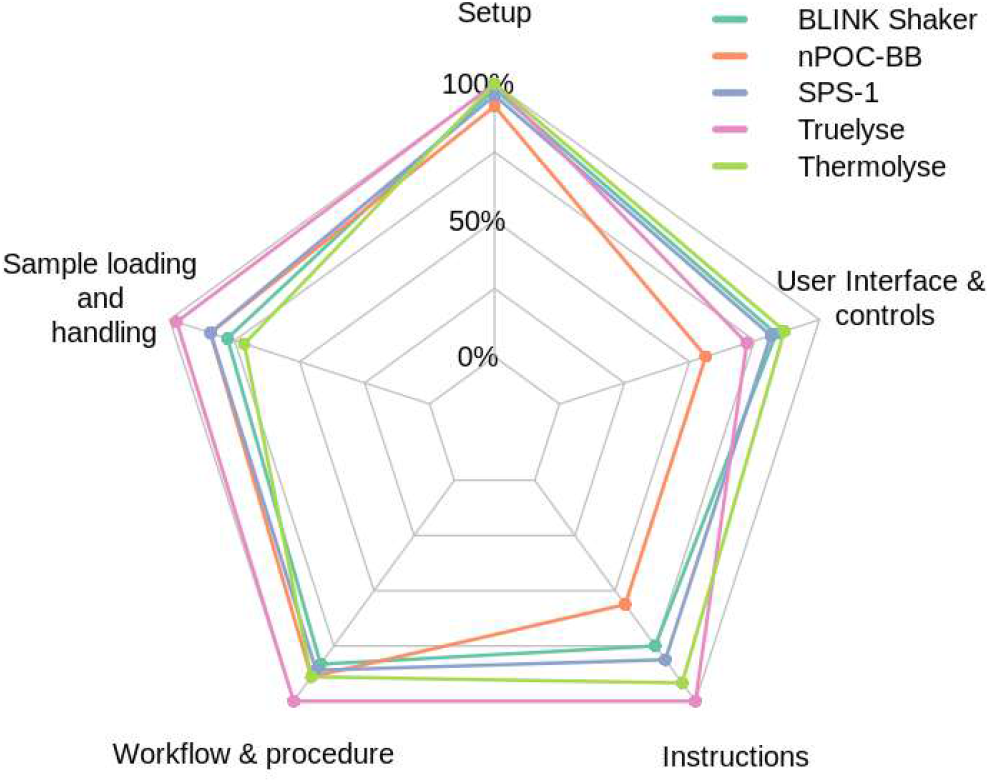
Radar plot comparing the usability scores of the test lysis devices across five predefined domains. Each axis represents the mean score for a single usability domain, averaged across three independent laboratory users who evaluated the devices using a standardized five-point Likert scale. Higher scores indicate greater usability.

## Discussion

This study provides independent analytical benchmarking of five novel, portable lysis devices across multiple performance domains, offering a reference framework against which future device development can be evaluated. Overall, lysis efficiency was strongly influenced by both buffer formulation and sample matrix, with most devices approaching benchmark performance under specific conditions.

Lysis efficiency varied considerably across devices, matrices, and buffer conditions. Bead-beating-based devices achieved recovery comparable to the reference workflow across MTB inputs in both tongue and sputum swabs when used with the reference buffer. Sonication-based devices exhibited matrix- and buffer-dependent performance, with recovery approaching or exceeding the reference workflow in the reference buffer, but reduced efficiency in tongue swabs when proprietary buffers were used. Heated-vortex-based lysis demonstrated comparatively lower DNA recovery across matrices and buffers. Overall, although most devices approached benchmark efficiency under specific conditions, performance was strongly influenced by both buffer formulation and sample matrix. A key observation across platforms utilising proprietary buffers was frothing during bead-beating, likely attributable to detergent content.

While detergents facilitate membrane disruption and fluid flow, excessive foaming may interfere with mechanical lysis, and optimising detergent concentration represents one avenue for improving performance. Similarly, the matrix-dependent variability observed in sonication-based devices highlights the need for matrix-specific optimisation: sputum contains higher bacterial loads alongside mucus and inhibitors, whereas tongue swabs reflect the oral environment, characterised by host epithelial cells and salivary enzymes, presenting distinct lysis challenges.^14–16^ In several instances, novel workflows exceeded reference DNA recovery, which may reflect improved nuclease inactivation when thermal and mechanical disruption occur simultaneously. Sequential workflows, heat followed by mechanical lysis, may allow delayed release of active nucleases, increasing the risk of DNA degradation before detection.

While this mechanistic relationship remains speculative, it provides a biologically plausible rationale for observed performance differences and warrants further investigation, for example through time-course experiments comparing simultaneous versus sequential lysis conditions.

A notable finding is the comparatively reduced lysis efficiency observed for the vortex-based device, which contrasts with the strong overall clinical performance of the MiniDock MTB Test (Guangzhou Pluslife Biotech, China), an integrated workflow incorporating this lysis mechanism that has received WHO recommendation.^2^ This apparent disconnect may partly reflect that in moderate-to high-burden disease, bacterial loads are sufficient to compensate for suboptimal lysis, masking its impact on diagnostic sensitivity. However, the MiniDock MTB Test has shown decreasing concordance with Xpert MTB/RIF Ultra (Cepheid, USA) across semiquantitative bacterial load grades, with the greatest discordance at very low and trace-positive levels, precisely the range where lysis efficiency is most likely to be limiting.^17^ This suggests that optimising lysis efficiency could directly address the sensitivity gap observed in paucibacillary samples, including those from patients with early disease, children, or HIV-associated TB. More broadly, this highlights an important role for analytical benchmarking: not as a direct predictor of clinical utility, but as a tool for identifying targeted optimisation opportunities in the patient populations most at risk of missed diagnosis.

Biosafety is a fundamental requirement for TB diagnostics at or near the point of care, with WHO guidance stipulating that sample processing risk should not exceed that of smear microscopy.^18^ Complete inactivation was achieved by bead-beating-based devices and the heated-vortex-based platform, whereas sonication-only devices showed residual bacterial growth. This suggests that sonication alone may not generate sufficient thermal energy for complete MTB inactivation, a finding with direct implications for device design. Integration of a thermal inactivation step is likely essential for sonication-based platforms to meet biosafety requirements, particularly in near-patient settings.

This study has several strengths. It provides independent, multi-domain benchmarking of five lysis devices, encompassing DNA recovery, nuclease inactivation, biosafety, and operational usability, against a well-characterized reference workflow. Evaluating both tongue and sputum swabs contributes comparative data to the rapidly expanding evidence base for swab-based TB diagnostics, a field where standardised analytical frameworks have been lacking.

Limitations should be noted as well. Nuclease inactivation was assessed in tongue swabs only. Equivalent assessment in sputum swabs is needed to fully characterize each device’s performance in that matrix. A formal Limit of Detection (LOD) assessment was not performed and represents a priority for future evaluation. Usability data were derived from a small cohort of skilled laboratory staff, limiting generalizability to real-world settings. Finally, several devices evaluated were prototypes, and performance should be reassessed following design-lock.

In summary, this study demonstrates that portable extraction-free lysis devices can achieve DNA recovery comparable to gold-standard laboratory methods, though performance is critically dependent on sample matrix, buffer composition, and lysis mechanisms. Analytical benchmarking of this kin is essential to guide device optimisation, particularly for paucibacillary samples where improvements in lysis efficiency are most likely to enhance diagnostic sensitivity and reduce missed diagnoses in vulnerable populations. These findings provide a multi-domain analytical framework to inform the next generation of simplified, field-deployable molecular TB diagnostic workflows.

## Declarations

### Ethics statement

Ethical approval was obtained by Global Health Labs from the institutional review board at WCH Clinical (20181925). Written informed consent was obtained from all volunteers. In accordance with institutional guidelines, ethical review was not sought from Heidelberg University Hospital, as the study did not meet the criteria for human subjects research requiring such approval.

### Contributions (CRediT)

Sonal Jain: Methodology; Investigation; Data curation; Formal analysis; Visualisation; Writing – original draft; Writing – review and editing.

Alexey Ball: Methodology; Investigation; Data curation; Formal analysis; Writing – review and editing

Caitlin Anderson: Methodology; Investigation; Writing – review and editing

Adithya Cattamanchi: Supervision; Writing – review and editing.

Claudia Denkinger: Supervision; Writing – review and editing.

Amy Steadman: Conceptualisation; Methodology; Supervision; Writing – review and editing.

Seda Yerlikaya: Conceptualisation; Methodology; Supervision; Writing – review and editing.

## Data availability

The data generated in this study are available from the corresponding author upon reasonable request.

